# “Kankasha” in Kassala: a prospective observational cohort study of the clinical characteristics, epidemiology, genetic origin, and chronic impact of the 2018 epidemic of Chikungunya virus infection in Kassala, Sudan

**DOI:** 10.1101/2020.09.23.20199976

**Authors:** Hilary Bower, Mubarak el Karsany, Abd Alhadi Adam Hussein, Mubarak Ibrahim Idriss, Ma’aaza Abasher AlZain, Mohamed Elamin Ahmed Alfakiyousif, Rehab Mohamed, Iman Mahmoud, Omer Albadri, Suha Abdulaziz Alnour Mahmoud, Orwa Ibrahim Abdalla, Mawahib Eldigail, Nuha Elagib, Ulrike Arnold, Bernardo Gutierrez, Oliver G. Pybus, Daniel P. Carter, Steven T. Pullan, Shevin T. Jacob, Tajeldin Mohammedein Abdallah, Benedict Gannon, Tom E. Fletcher

**Affiliations:** UK Public Health Rapid Support Team, London School of Hygiene & Tropical Medicine/Public Health England, London, United Kingdom; National Public Health Laboratory, Federal Ministry of Health, Khartoum, Sudan; Karary University, Omdurman, Sudan; University of Kassala, Kassala, Sudan; Laboratory Division, Kassala State Ministry of Health, Kassala, Sudan; Communicable Disease Surveillance & Events Unit, Federal Ministry of Health, Khartoum, Sudan; Health Emergency and Epidemic Control Directorate, Federal Ministry of Health, Khartoum, Sudan; Department of Zoology, University of Oxford, Oxford, United Kingdom; National Infection Service, Public Health England, Porton, United Kingdom; Kassala Teaching Hospital, Kassala, Sudan; Clinical Sciences, Liverpool School of Tropical Medicine, Liverpool, United Kingdom

**Keywords:** Chikungunya, disability, epidemiology, outbreak, Kassala, Sudan

## Abstract

**Background:** The public health impact of Chikungunya virus (CHIKV) is often underestimated. Usually considered a mild condition of short duration, recent outbreaks have reported greater incidence of severe illness, fatality, and longer-term disability. In 2018/19, Eastern Sudan experienced the largest epidemic of CHIKV in Africa to date, affecting an estimated 487,600 people. Known locally as Kankasha, this study examines the clinical characteristics, risk factors, and phylogenetics of the CHIKV epidemic in Kassala City.

**Methodology/Principal Findings:** A prospective cohort of 142 cases (102 adults, 40 children) were enrolled at Kassala Teaching Hospital in October 2018. Clinical information, socio-demographic data and sera samples were analysed to confirm diagnosis, characterise illness, and identify the viral strain. CHIKV infection was confirmed by real-time reverse transcription-PCR in 84.5% (120/142) of participants. Nine had concurrent CHIKV/Dengue virus (DENV) infection and 28.8% had a positive Rapid Diagnostic Test for malaria. Five percent had haemorrhagic symptoms including two children with life-threatening haemorrhage. One CHIKV-positive participant died with acute renal injury.

Ninety to 120 days post-illness, 63% of those followed-up were still experiencing arthralgia in one or more joints, and 11% remained moderately disabled using Rapid3 assessment. Phylogenetic analysis showed all CHIKV infections belonged to a single clade within the Indian Ocean Lineage (IOL) of the East/Central/South African (ECSA) genotype. History of contact with an infected person was the only socio-demographic factor associated with infection (p=0.01), suggesting that vector transmission in households is important.

**Conclusions/Significance:** The epidemic is estimated to have affected ∼ 50% of Kassala City’s population. Substantial vulnerability to CHIKV remains here and elsewhere in Sudan due to widespread *Aedes aegypti* presence and mosquito-fostering household water storage methods. This study highlights the importance of increasing awareness of the severity and socio-economic impact of CHIKV outbreaks and the need for urgent actions to reduce transmission risk in households.

**Author summary:** Chikungunya is an arboviral disease transmitted to humans by infected mosquitoes and characterised by fever and arthralgia. Although it is generally considered a short self-limiting infection, long term sequelae and severe disease are increasingly recognised. In 2018/19, Eastern Sudan experienced the largest epidemic of Chikungunya in Africa to date, affecting approximately 500,000 people. We undertook a prospective hospital-based cohort study of patients presenting with undifferentiated febrile illness in Kassala city, Sudan, supported by next-generation sequencing. We confirmed that CHIKV was the dominant pathogen, with positive CHIKV RT-PCR in 85% of patients presenting during the 7-day study period. Dengue virus was also circulating with nine CHIKV PCR-positive patients co-infected, and we identified high rates of *Plasmodium falciparum* malaria infection and CHIKV/malaria co-infection. Genetic sequencing confirmed Indian Ocean Lineage of the East/Central/South African CHIKV genotype. A substantial proportion of participants were admitted to hospital including children with haemorrhage, reflecting the severe phenotype linked to this genotype. Increased understanding of the health and economic burden of Chikungunya is needed, and recognition that severe and occasionally fatal infection exists. With widespread presence of *Ae. aegypti* and household water storage practices that encourage mosquito breeding, timely actions will be essential to prevent further large outbreaks.

## INTRODUCTION

Chikungunya is often considered a mild illness and its public health impact underestimated in Africa and the Middle East despite the fact that greater severity and long-term sequelae have been reported increasingly in recent years.^1-3^

First isolated in Tanzania in 1952,^4^ Chikungunya virus (CHIKV) was implicated in rural outbreaks and sporadic cases across Africa until 1980, when it effectively disappeared. In 2004, however, it reappeared with a vengeance in a pandemic that spread from Kenya to the Indian Ocean islands and Asia, causing millions of infections.^5, 6^ In central Africa, sizeable CHIKV outbreaks in cities signalled a major shift in transmission to urban settings.^2, 3, 7-9^

Early symptoms of CHIKV are similar to many tropical febrile illnesses but it is differentiated by debilitating arthralgia, often bilaterally and in multiple joints. Other symptoms include headache, gastrointestinal problems, fatigue, asthenia, peripheral oedema and conjunctivitis.^10^ Although most cases improve after 1-2 weeks, recent outbreaks have seen higher incidences of severe illness, including sepsis and cardiac, renal, neurological, skin and ocular manifestations, and of longer-term effects including persistent pain, rheumatic symptoms, depression, and mood and sleep disorders.^11-16^ A meta-analysis of chronic symptoms studies in 15 outbreak countries found 43% of cases had not recovered at three months, and 21% unrecovered at 12 months. Long term neurodevelopmental delays have been reported in infants symptomatically infected through vertical transmission.^17, 18^ Apart from anti-inflammatory drugs, there is no treatment for Chikungunya and, as yet, no vaccine.^19^

Recent outbreaks have also challenged the view that CHIKV fatality is rare. Mortality of 10-48% were reported in the Reunion Island outbreak among cohorts of hospitalised patients.^13, 16^ Central nervous system Infections with fatal outcomes have been reported in the Americas.^20^ Other locations have documented substantial rises in all-cause deaths during outbreak periods.^20^

Hypotheses to explain these changes include persistent immune activation triggered by viral debris, improved surveillance and genetic mutation, particularly a new variant known as the Indian Ocean Lineage (IOL) which, since the 2004-2006 pandemic, has been linked to greater severity and the highest rates of non-recovery.^21^ IOL has also been linked to increased risk of outbreak as some forms contain a mutation that allows CHIKV to use the temperate-dwelling *Aedes albopictus* mosquito as a vector as well the tropical *Aedes aegypti*.^5, 21^

In low-resource settings, outbreaks of undifferentiated febrile illness (UFI) are both common and a diagnostic challenge. The Federal Republic of Sudan has seen 12 major outbreaks of UFI since 2012, frequently associated with haemorrhage and high case fatality ratios^22^ and a range of high-consequence infectious diseases are endemic or epidemic in Sudan. Crimean-Congo haemorrhagic fever was recently identified as a common pathogen in a retrospective investigation of outbreaks in Darfur.^23^ Yellow fever virus epidemics occur^24^ and outbreaks of dengue fever are common.^25^ The Federal Ministry of Health (FMoH) maintains a sentinel surveillance system and supports individual states with rapid response teams and enhanced diagnostic capability at the National Public Health Laboratory (NPHL) in Khartoum.

In late July 2018, the sentinel surveillance in Kassala State (Figure 1) detected an increased frequency of UFI and CHIKV was identified by the NPHL in the blood sample of a traveller to Kassala city from nearby Red Sea State. In mid-September reports of cases with more severe symptoms, including haemorrhage, raised fears that another pathogen might be involved. Using a pre-prepared, locally and internationally-approved protocol, FMoH and Kassala Teaching Hospital (KTH), supported by Kassala University and the UK Public Health Rapid Support Team, deployed a small pre-trained study team of clinical, epidemiology and laboratory staff to investigate the outbreak syndrome, confirm the outbreak pathogen, and sequence the outbreak strain.

**Figure 1:**
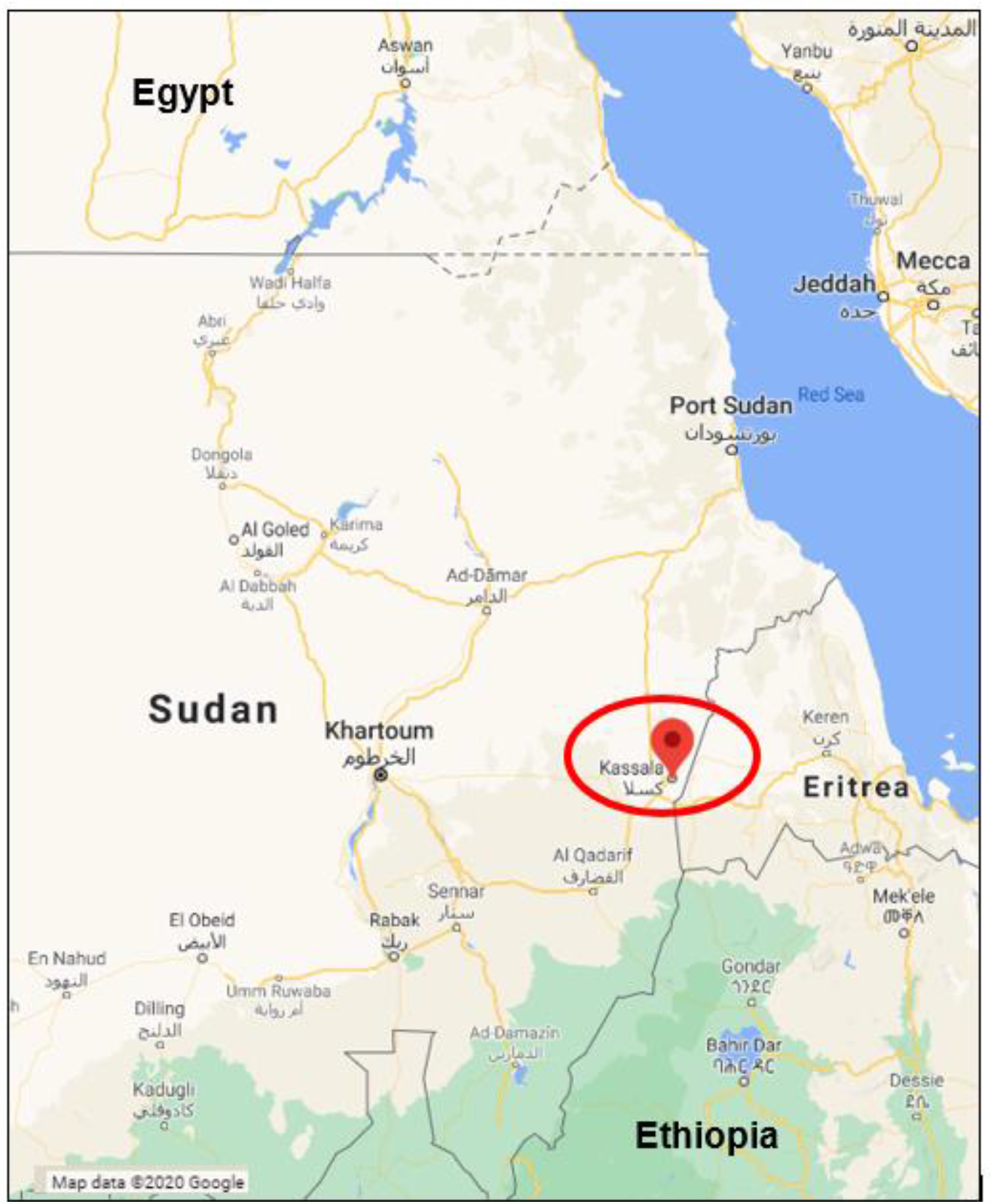
Study location. Legend: Kassala state, located in Eastern Sudan, 600 km the capital city, Khartoum. covers an area of 42,282 km^2^ and has a population 1.8 million inhabitants. Kassala City is the State capital with a population of ∼ 400,000. Kassala Teaching Hospital is the State tertiary hospital and provides service for all patients referred from health centres and rural hospitals.

## METHODS

The study was a prospective hospital-based observational cohort of consecutive patients presenting with UFI (case definition, Figure 2) at the KTH medical and paediatrics outpatient clinics departments between 10th and 16th October 2018. Epidemiological information, clinical symptoms and laboratory results were recorded at presentation on standardised pro-formas (Supporting Information 1). The sample size of 140 cases was informed by existing literature3 and feasible recruitment in the urgent time frame (7 days).

**Figure 2:**
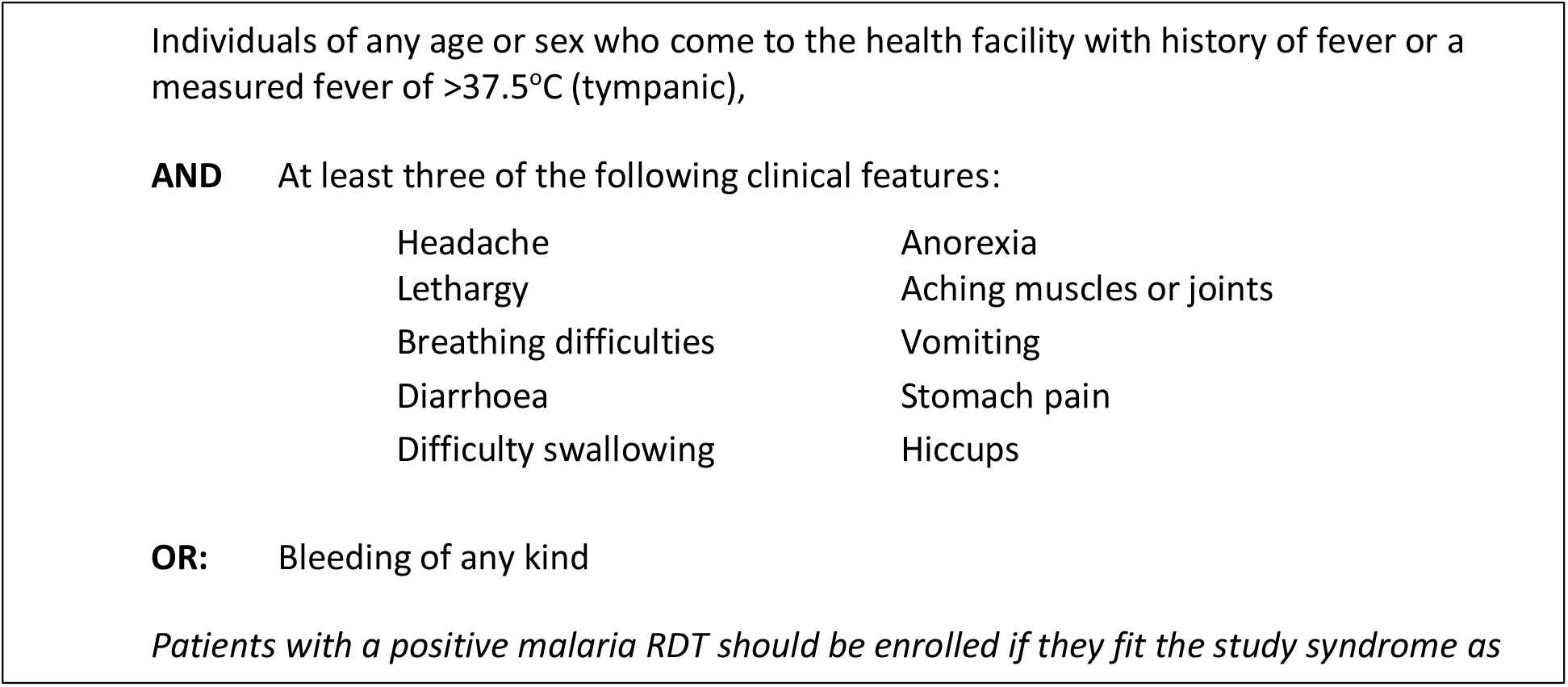
Case definition used in Kassala Teaching Hospital.

Blood samples were drawn at initial assessment, and adults who responded to phone follow-up provided convalescent samples and completed the WHO-validated Routine Assessment of Patient Index Data 3 (RAPID3) disability and pain survey^26^ 90-120 days later (Jan-Feb 2019). Children were not asked to return due to common reluctance to allow blood-draw from healthy children, but outcome and duration of hospital stay were confirmed with parents. All contactable participants were given their PCR test results.

Haematology (Mindaray 3000 Plus, China), biochemistry analysis (Biosystem BTS 310, Germany), and malaria rapid diagnostic tests (RDT, SD Bioline, USA) were performed at admission on all participants: malaria thick film examination was done for all children. Plasma samples were frozen at -80°C and transported under liquid nitrogen to the NPHL in Khartoum where nucleic acid was extracted (QIAamp Viral RNA Mini kit, Qiagen, Germany) and qRT-PCR performed to detect CHIKV RNA (RealStar Chikungunya RT-PCR Kit 2.0, Altona Diagnostics, Germany) using Rotor Gene Q or Corbett RG6000 thermocyclers. Exposure to CHIKV and DENV was assayed by anti-chikungunya and anti-dengue IgM and IgG indirect ELISA (Euroimmun, Germany). Due to limited availability of anti-chikungunya IgM ELISA kits, CHIKV PCR-negative samples were prioritised, followed by 69 CHIKV PCR-positives in order of recruitment. An Ebolavirus qRT-PCR (RealStar Ebolavirus RT-PCR Kit 1.0, Altona Diagnostics, Germany) was performed on samples from patients with bleeding symptoms.

All patient data were entered into encrypted software and analysed with STATA (StataCorp LLC, USA, V14.2). Descriptive statistics are expressed as medians and Inter-Quartile Range (IQR) or means and standard deviations (SD) for continuous variables, and frequencies and proportions for categorical variables. Fisher’s exact, Chi^2^, T-tests and Spearman’s Rank Order coefficients were used to assess association and correlation related to CHIKV PCR result. Significance was set at p<0.05.

Ethical approval was granted by the FMoH Technical Review Board, and Ethics Committee of Karary University, Khartoum and the London School of Hygiene & Tropical Medicine (Ref: 11930). All participants or parents/guardians of children aged under 18 years provided written informed consent.

### Genetic sequencing

Complementary DNA (cDNA) was prepared and Sequence Independent Single Primer Amplification (SISPA) performed prior to library preparation using Oxford Nanopore kits (SQK-LSK108). Sequencing on flow cells (FLO-MIN106) was done using the MinION (Oxford Nanopore Technologies) as previously described.^27^ Thirty samples were sequenced at NPHL and analysed using Albacore 1.2 to basecall, Mash Screen^28^ was used to identify the most closely matched genome on Genbank and a reference guided genome assembly constructed as described previously.^27^ The first seven sequences were used to construct a basic phylogenetic analysis using MEGA version 6 (results not shown) in order to identify the Kassala strain in Sudan. With FMoH permission, sample aliquots were also transferred to PHE Porton and sequenced using the same methods. The resulting viral genetic sequences were sent to Oxford University Department of Zoology for phylogenetic analysis.

Whole genome sequences were aligned with publicly available complete CHIKV genomes belonging to the ECSA genotype using MAFFT *v7*.*450* implemented in Geneious R8.^29^ A maximum likelihood (ML) phylogenetic tree was estimated using RAxML 8.0^30^ under a GTR substitution model with a Gamma model of among-site rate heterogeneity. The estimated ML phylogeny was midpoint rooted, and node support was assessed through 100 ML bootstrap replicates. The geographic location and amino acid identity at site E1-226 of each sequence were annotated in the tree to explore the phylogenetic distribution of those traits.

## RESULTS

A total of 102 adults and 42 children (<18 years) were recruited over seven days, of which two cases without samples were excluded. 11 patients or their guardians refused to participate, and 2 patients dead on arrival were not included as sampling was not possible (Figure 3).

**Figure 3:**
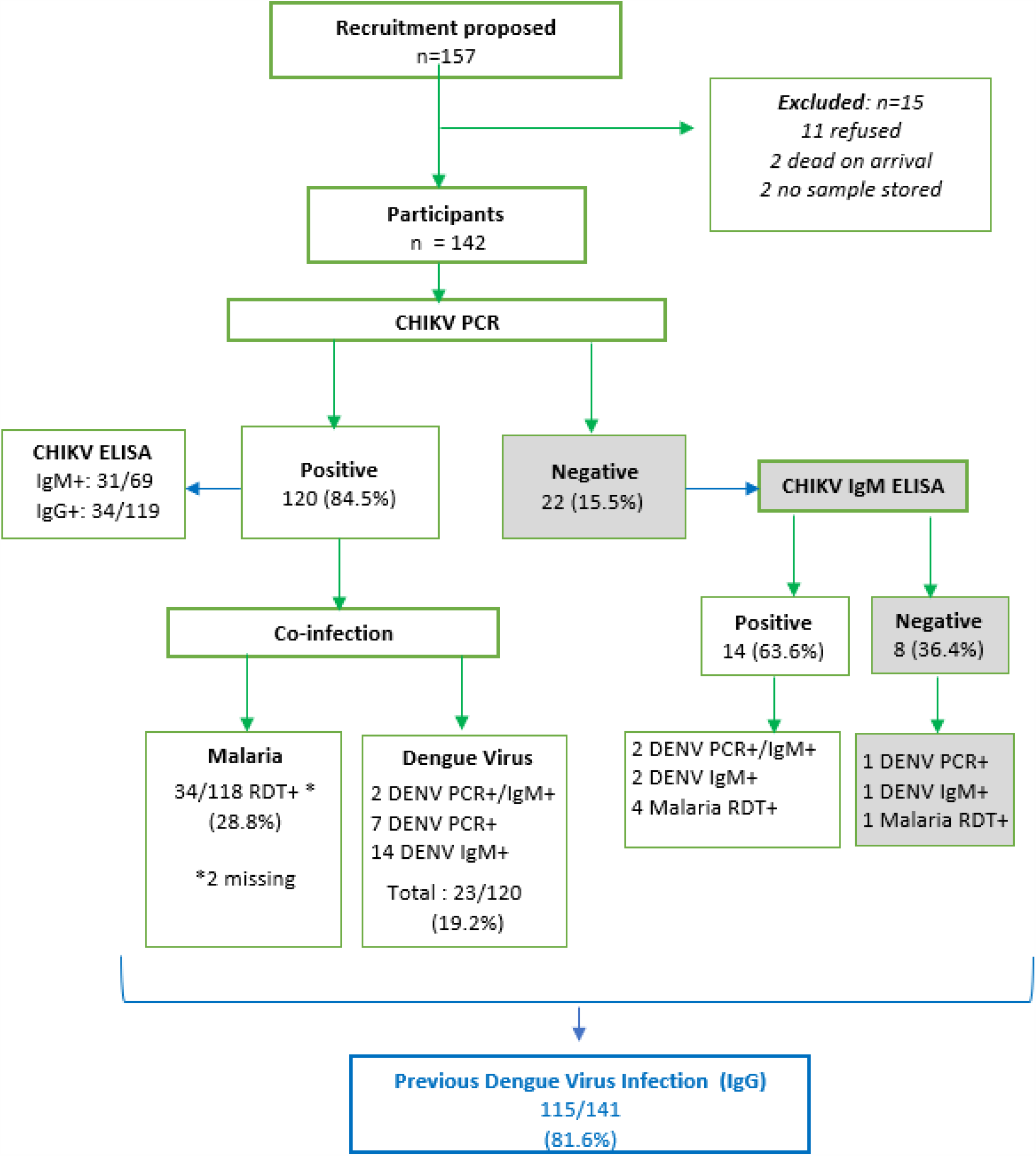
Flow diagram of study participants and virological findings.

Participants were aged 4 months to 70 years (mean 27 years, SD 17.6); 47.9% were female, none were pregnant (Table 1). Almost half (46.1%, 65/141) were people likely to spend more time in a household compound (e.g. housewives, unemployed, retired, children under 5). Median household size was 8. Two-thirds of participants lived in brick/concrete houses with own well and sanitation, the remainder lived in less permanent material structures with shared water and sanitation. A third kept animals in their compound.

**Table 1:**
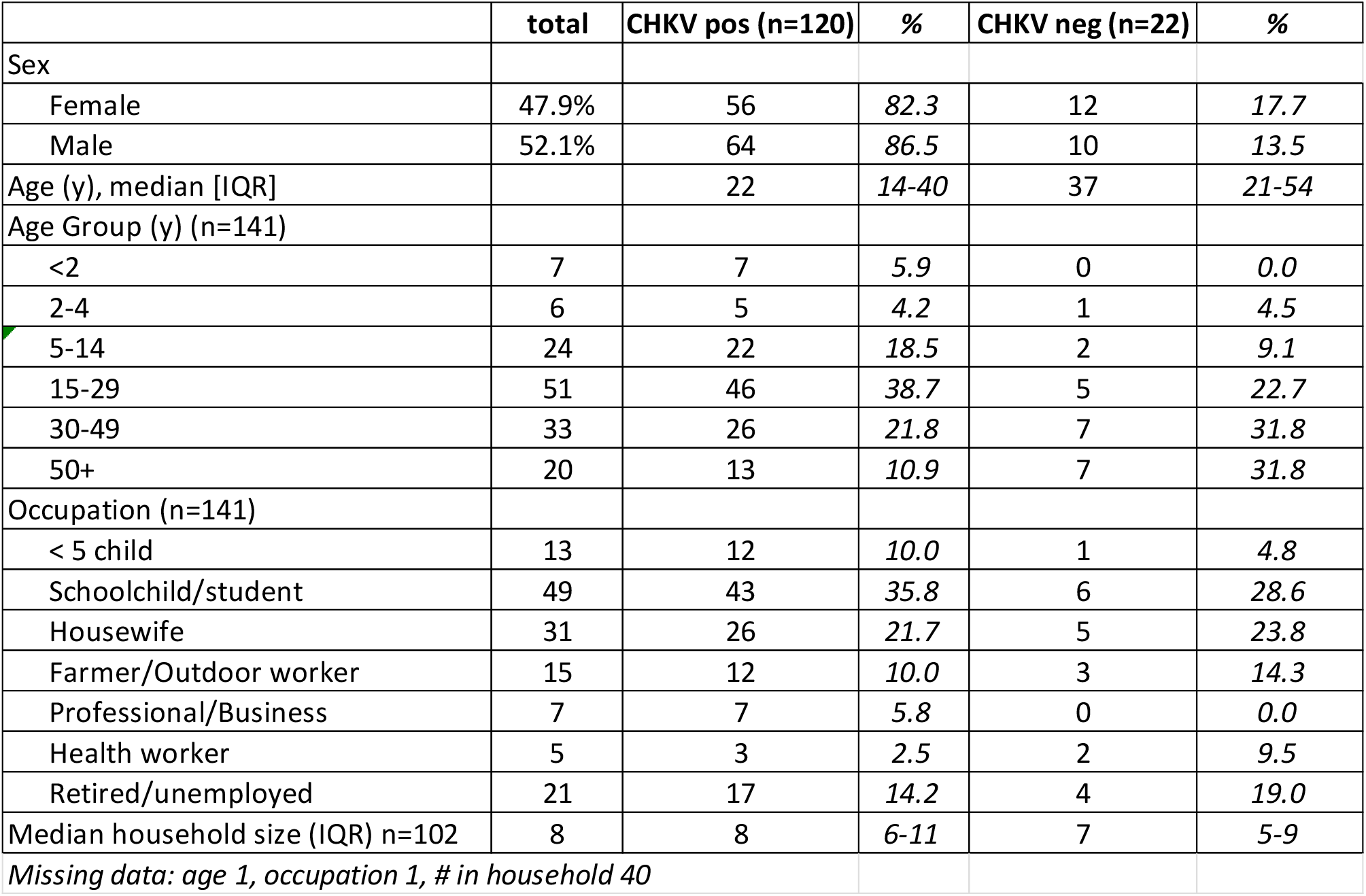
Participant characteristics by Chikungunya qRT-PCR result.

|  | total | CHKV pos (n=120) | % | CHKV neg (n=22) | % |
| --- | --- | --- | --- | --- | --- |
| Sex |  |  |  |  |  |
| Female | 47.9% | 56 | 82.3 | 12 | 17.7 |
| Male | 52.1% | 64 | 86.5 | 10 | 13.5 |
| Age (y), median [IQR] |  | 22 | 14-40 | 37 | 21-54 |
| Age Group (y) (n=141) |  |  |  |  |  |
| <2 | 7 | 7 | 5.9 | 0 | 0.0 |
| 2-4 | 6 | 5 | 4.2 | 1 | 4.5 |
| 5-14 | 24 | 22 | 18.5 | 2 | 9.1 |
| 15-29 | 51 | 46 | 38.7 | 5 | 22.7 |
| 30-49 | 33 | 26 | 21.8 | 7 | 31.8 |
| 50+ | 20 | 13 | 10.9 | 7 | 31.8 |
| Occupation (n=141) |  |  |  |  |  |
| < 5 child | 13 | 12 | 10.0 | 1 | 4.8 |
| Schoolchild/student | 49 | 43 | 35.8 | 6 | 28.6 |
| Housewife | 31 | 26 | 21.7 | 5 | 23.8 |
| Farmer/Outdoor worker | 15 | 12 | 10.0 | 3 | 14.3 |
| Professional/Business | 7 | 7 | 5.8 | 0 | 0.0 |
| Health worker | 5 | 3 | 2.5 | 2 | 9.5 |
| Retired/unemployed | 21 | 17 | 14.2 | 4 | 19.0 |
| Median household size (IQR) n=102 | 8 | 8 | 6-11 | 7 | 5-9 |
| Missing data: age 1, occupation 1, # in household 40 |  |  |  |  |  |

Most participants came from Kassala City sectors 2,3, 4 and 5 on the banks of the seasonal River Gash (69%, 80/116), coinciding with areas of greatest flooding in the 2018 rainy season, and with highest case reports during the epidemic (Figure 4). A further 26% (30/116) were from rural areas up to 1.5 hours’ drive away.

**Figure 4:**
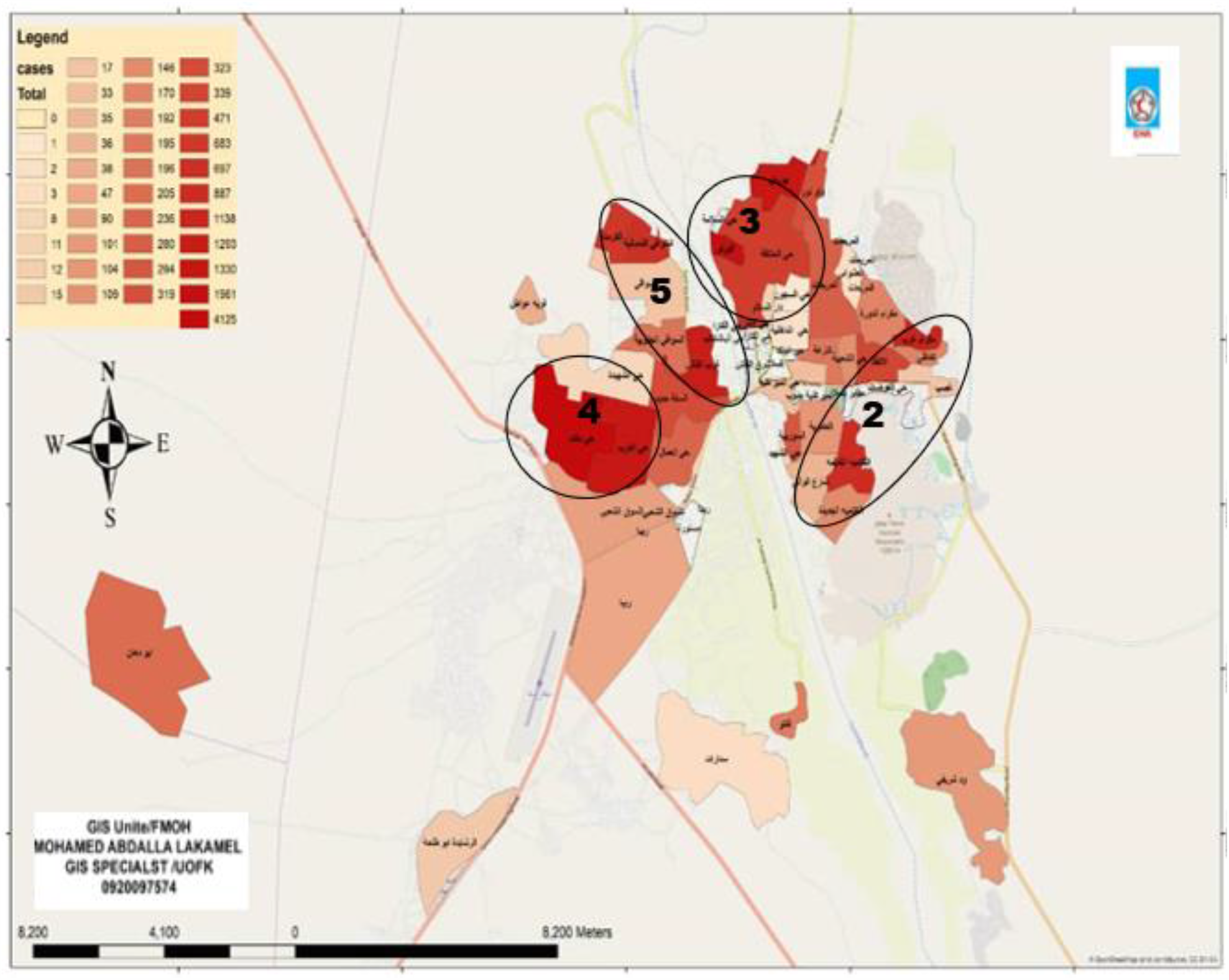
Distribution of all reported Chikungunya cases in Kassala City and surrounds, Aug – Nov 2018 (n=19,902) Credit: Kassala State Ministry of Health Epidemiology Unit, Epidemic Report Presentation, 5 Nov. 2018 Circles represent City Sectors 2,3,4,& 5

### Clinical presentation

Median delay from symptom onset to presentation was 2 days (IQR 1-4, n=139) with no difference by CHIKV diagnosis or age (Table 2). Sixteen (26.6%) participants were admitted (4 adults, 12 children). Children were more likely to be admitted than adults and for longer: a mean of 4.7 days compared to 2.3 days for adults (p=0.04).

**Table 2:** Clinical symptoms and Ct values of participants at presentation by CHIKV qRT-PCR result (n=142)

|  | CHIKV pos | % | CHIKV neg | % |
| --- | --- | --- | --- | --- |
| **Days from onset to presentation** | 2 | *IQR 1-4* | 2 | *IQR 1-5* |
| **Symptoms at presentation** |  |  |  |  |
| Fever/history of fever | 117 | 97.5 | 22 | 100.0 |
| Any bleeding | 6 | 5.0 | 1 | 4.8 |
| Headache | 97 | 80.8 | 17 | 85.0 |
| Joint pain | 93 | 77.5 | 15 | 75.0 |
| Fatigue | 85 | 70.8 | 17 | 81.0 |
| Muscle pain | 66 | 55.0 | 16 | 76.2 |
| Back pain | 55 | 45.8 | 9 | 42.9 |
| Loss of appetite | 46 | 38.3 | 6 | 28.6 |
| Vomiting | 45 | 37.5 | 8 | 38.1 |
| Abdominal pain | 32 | 26.7 | 8 | 38.1 |
| Lack of strength | 25 | 20.8 | 4 | 100.0 |
| Dizziness | 18 | 15.0 | 3 | 14.3 |
| Chest pain | 17 | 14.2 | 3 | 15.0 |
| Cough | 17 | 14.2 | 2 | 9.5 |
| Rash | 13 | 10.8 | 2 | 9.5 |
| Diarrhoea | 11 | 9.2 | 2 | 10.0 |
| Dysphagia | 9 | 7.5 | 3 | 15.0 |
| Dehydration | 9 | 7.5 | 1 | 33.3 |
| Shortness of breath | 7 | 5.8 | 2 | 10.0 |
| Conjunctivitis | 7 | 5.8 | 3 | 15.0 |
| Sore throat | 6 | 5.0 | 0 | 0.0 |
| Confusion | 3 | 2.5 | 0 | 0.0 |
| **PCR Cycle Threshold** |  |  |  |  |
| High (Ct <20) | 33 | 27.5 | - |  |
| Moderate (Ct 20-29) | 24 | 20.0 | - |  |
| Low (Ct 30-38) | 63 | 52.5 | - |  |
| **Other pathology** |  |  |  |  |
| Malaria positive (RDT) | 34/118 | 28.8 | 5/22 | 22.7 |
| DENV positive (PCR) | 9/120 | 7.5 | 3/22 | 13.6 |
| *\*Missing: onset date 3; malaria RDT 2;* |  |  |  |  |

Most common symptoms at presentation among CHIKV PCR-positive participants were fever (97.5%), headache (88.2%), fatigue (82.5%), muscle, joint and back pain (66.0%, 83.8%, 50.9% respectively), loss of appetite (40.4%) and vomiting (40.2%); patients who were CHIKV PCR-negative had similar presenting symptoms. Participants co-infected with CHIKV and DENV were more likely to present with back pain than those with CHIKV alone (p=0.003). Five percent (6/120) of CHIKV PCR-positive participants reported bleeding, including haematemesis (4), oral bleeding (2), epistaxis (3), petechiae (1), haemoptysis (1) and melaena (1). Ten percent of CHIKV PCR-positive adults (18/78) were hypotensive (systolic blood pressure <100 mmHg) with seven also being tachycardic (pulse >100).

### Laboratory results

Of 142 participants, 120 were confirmed positive for CHIKV infection by qRT-PCR, giving the case definition used a positive predictive value of 84.5%. CHIKV PCR cycle threshold (Ct) values ranged from 12-37 (mean 27.3, SD 7.6); over a quarter (27.5%) had a Ct value corresponding with a high viral load (Ct<20) (Table 2). A significant correlation existed between Ct value and both day of illness (Spearman r=0.37, p<0.001) and lymphocyte count (r=0.23, p=0.012); no other significant correlations were found. Prolonged viremia was observed in seven CHIKV PCR-positive who presented 10 days after disease onset. Of 91 participants tested for CHIKV IgM antibodies, 45 were positive including 14 participants who were negative on CHIKV PCR (Figure 3).

Nineteen percent of CHIKV PCR-positive participants (23/120) were coinfected with DENV either by PCR or IgM-ELISA. Of all samples (CHIKV-positive and negative), 81.6% (115/141) were DENV IgG-positive indicating previous infection. Of the 118 CHIKV PCR-positive participants with malaria RDT result, 28.8% were positive, with the highest proportion in those aged 15-29 years. No participants were co-infected with all three pathogens.

Among participants with matched CHIKV PCR and ELISA results, 31/69 PCR-positive were also IgM-positive, and 34/119 (28.6%) were CHIKV IgG positive (Figure 3). Participants who were CHIKV positive on both PCR and IgM presented later in illness course than those who were CHIKV PCR-positive and IgM-negative (p<0.001). This finding was replicated with CHIKV PCR & IgG-positive patients who presented later than CHIKV IgG-negative patients (p=0.002).

There was a significant difference in mean haematocrit levels between CHIKV PCR-positive (mean 35.9, SD=6.27) and negative participants (mean 38.9, SD=7.41, p=0.05), but no other significant differences in blood chemistry. Heart rate and blood pressure were similar in both groups. In the CHIKV PCR-positive cohort median haematology and biochemistry results were all within normal ranges (Table 3). Leucopenia (white blood count < 4 × 10^9^/L) was observed in 29% (35/120) and lymphopenia (< 1 × 10^9^/L) observed in 50.7% (61/120). Elevated AST (>40IU/L) was observed in 22.5% (27/120) with six participants at levels> 100 IU/L (max 259 IU/L). ALT levels >51 IU/L were seen in seven participants with four recording levels >100 (max 150 IU/l). Acute kidney injury was observed in one fatal case. Platelet counts < 100 × 10^9^/L were recorded in 23/120 CHIKV PCR-positive participants, of whom 14/23 were also malaria RDT positive and none DENV PCR positive. Platelet counts < 50 × 10^9^/L were observed in six participants, three of whom also had positive malaria RDT.

**Table 3:** Mean clinical parameters of participants at presentation by CHIKV qRT-PCR result.

|  | CHIKV+ | SD | *n* | CHIKV- | SD | *n* |
| --- | --- | --- | --- | --- | --- | --- |
| Heart rate (beats/min) | **91.2** | 19.2 | 114 | **83.5** | 15.8 | 21 |
| Respiratory rate | **20.4** | 2.9 | 12 | **18.0** | n/a | 1 |
| Systolic pressure | **102.9** | 20.2 | 78 | **106.3** | 12.5 | 20 |
| Diastolic pressure | **68.0** | 13.1 | 78 | **70.2** | 7.5 | 20 |
| Haemoglobin (g/dL) | **11.6** | 2.2 | 120 | **12.5** | 2.5 | 22 |
| Haematocrit (%)<sup>*</sup> | **35.9** | 6.3 | 119 | **38.9** | 7.4 | 22 |
| WBC (10<sup>3</sup>/uL) | **6.1** | 3.1 | 120 | **4.8** | 2.2 | 22 |
| Lymphocytes (10<sup>3</sup>/uL) | **2.1** | 2.4 | 118 | **2.9** | 6.6 | 22 |
| Platelets (10<sup>9</sup>/L) | **227.0** | 143.3 | 120 | **190.6** | 131.0 | 22 |
| Nitrogen oxide | **3.8** | 2.0 | 113 | **2.9** | 1.7 | 22 |
| Urea (mg/dL) | **24.8** | 14.9 | 119 | **26.1** | 11.3 | 22 |
| Creatinine (mg/dL) | **0.8** | 0.7 | 119 | **0.9** | 0.2 | 22 |
| Bilirubin | **0.7** | 1.5 | 119 | **0.9** | 1.3 | 22 |
| AST (IU/L) | **36.0** | 33.9 | 119 | **28.2** | 17.4 | 22 |
| ALT (IU/L) | **23.1** | 23.4 | 118 | **21.5** | 14.4 | 22 |
| Albumin | **14.9** | 6.8 | 69 | **17.6** | 6.4 | 17 |
| *Significant difference: p=0.049 |  |  |  |  |  |  |

### Severe or fatal disease

The study recruited one case with a fatal outcome - a male teacher aged 52 transferred to KTH 13 days after onset of symptoms. He was CHIKV PCR-positive with a Ct of 30 and no coinfection. At admission he had petechiae and minor bruising but no other evidence of bleeding. He died from sepsis complicated by disseminated intravascular coagulation and acute kidney injury (Blood/Urea/Nitrogen (BUN) 154 mg/dL, creatinine 7.8 mg/dL, AST 259 IU/L, ALT 59 IU/L, platelets 111 × 10^9^/L).

Two children *in extremis* with significant haemorrhage were recruited at the paediatric department. Both were CHIKV PCR-positive without coinfection. The 16-year-old was admitted on day 2 of illness with a history of fatigue, headache, back pain, anorexia, oral mucosal haemorrhage, and profuse epistaxis. He was in shock (pulse 120, BP 84/50, respiratory rate 25) with a platelet count of 10 × 10^9^/L, Hb 12.5 g/dL, WBC 10.5 × 10^3^/uL, HCT 38.9%, AST 47 IU/L, ALT 24 IU/L, BUN 26 mg/dL, creatinine 0.8 mg/dL and CHIKV PCR Ct 29.63. He was resuscitated with blood and platelet transfusion, underwent nasal packing, and was discharged after 15 days. The second child aged 7 years presented on day 0 of illness with epistaxis, haematemesis, melaena, bruising and anorexia. Her pulse was 130 and Hb 10.2 g/dL, WBC 4.6 × 10^3^/uL, platelet count 20 × 10^9^/L, HCT 30.7%, urea 22 mg/dL, creatinine 0.5 mg/dL, AST 175 IU/L, ALT 129 IU/L and CHIKV PCR Ct of 26.8. She was also resuscitated with blood and platelets and discharged well after nine days admission.

### Factors associated with CHIKV positivity

The mean age of CHIKV PCR-positive patients was significantly less than CHIKV PCR-negative patients (p=0.003). CHIKV PCR-positives were also more likely to report exposure to someone who was ill (p=0.01) but as CHIKV is not transmitted person-to-person, this finding likely relates to being in the same vector environment. No associations were found with any other exposure. Compared to those with a single infection, CHIKV/DENV coinfected participants had higher mean systolic blood pressure (p=0.03) haemoglobin (p=0.01), haematocrit (p=0.03), bilirubin (p=0.002) and albumin (p=<0.001).

### Convalescent follow-up

Thirty (29.7%) of the 102 adult participants could be followed-up 90-120 days after enrolment, 29 (28.7%) refused, and 42 (41.6%) could not be reached. Parents of 30 of the 40 child participants responded to contact. All 60 followed-up were CHIKV PCR-positive at baseline. Sixteen (26.6%) had been admitted to hospital of whom four were children admitted for more than seven days (Table 4). Respondents were more likely to be admitted if they had bleeding (p=0.016), vomiting (p=0.014), or a positive malaria RDT (p=0.001). Of adults followed-up 43.3% (13/30) had spent 1-2 weeks off work and 20.0% (6/30) 2 or more weeks. Twenty-three of the 30 convalescent samples collected (76.7%) were IgG positive; 13 had seroconverted since recruitment and seven remained IgG negative.

**Table 4:** Impact of CHIKV infection on work and disability 90-120 days after acute infection.

|  | Adults (n=101) | %/SD | Children (n=40) | %/sd | p (exact) |
| --- | --- | --- | --- | --- | --- |
| **Follow-up response rate** | 30 | 29.7% | 30 | 75.0% |  |
| **Hospital care (n=30)** |  |  |  |  |  |
| Discharged the same day | 26 | 86.7% | 18 | 60.0% | p=0.039 |
| Admitted for at least 1 day | 4 | 13.3% | 12 | 40.0% |  |
| Mean days admitted | 2.3 | sd 1.3 | 4.7 | sd 4.1 |  |
| **Absence from work** |  |  |  |  |  |
| < 1 week | 11 | 36.7% |  |  |  |
| 1-2 weeks | 13 | 43.3% |  |  |  |
| 3-4 weeks | 4 | 13.3% |  |  |  |
| >4 weeks | 2 | 6.7% |  |  |  |
| **RAPID3 disability score (n=28)** |  |  |  |  |  |
| Near remission | 8 | 28.6% |  |  |  |
| Low disability | 17 | 60.7% |  |  |  |
| Moderate disability | 3 | 10.7% |  |  |  |
| **Current presence of pain (n=30)** | 19 | 63.3% |  |  |  |
| **Location of pain (multiple possible )** |  |  |  |  |  |
| Knee | 14 | 46.7% |  |  |  |
| Ankle | 11 | 36.7% |  |  |  |
| Wrist | 9 | 30.0% |  |  |  |
| Shoulder | 8 | 26.7% |  |  |  |
| Fingers | 7 | 23.3% |  |  |  |
| **Median pain perception score** | 4 | IQR 2-5 |  |  |  |
| **Median wellbeing score** | 7 | IQR 3-8 |  |  |  |

Note: Pain perception and wellbeing are scored on a 0-10 scale with 10 being the worst pain and feeling the worst
[Data access link to be added]

### Chronic disability

Nineteen of the 30 adults followed up at 90-120 days (63%) were still experiencing arthralgia, most frequently in the knee (46.7% 14/30), ankle (36.7% 11/30) wrist (30.0% 9/30) and shoulder (26.7% 8/30). Asked to rank pain and general wellbeing over the previous week on a scale where 10 was the most pain/most unwell, respondents reported a median pain score of 4 (IQR 2-5) and median wellbeing score of 7 (IQR 3-8). A third (36.6%, 11/30) indicated their pain was greater or the same as during the acute phase of their illness, and even those with less pain than before reported continuing to take painkillers, anti-inflammatories, corticosteroids, or all three, for their condition. In total, 60% continued to take medication.

Of the 28 respondents who completed the RAPID3 disability survey, three-quarters reported a continuing effect on day-to-day activities, including difficulties getting out of bed (28%), walking 2 miles (35%) and getting in and out of a vehicle (17%), while 10-15% reported difficulty with activities such as turning on taps, bending and sleeping well. Rapid3 scaling indicated that, 90-120 days after their acute illness, 10.7% (3/28) of respondents were experiencing moderately severe disability and 60.7% (17/28) low severity disability. The remaining eight (28.6%) respondents were considered nearly recovered. Neither level of viremia on admission, nor pre-existing conditions such as diabetes, hypertension and osteoarthritis, were associated with level of residual disability, or duration of absence from work.

### Genetic analysis

Virus genetic sequencing found that all CHIKV PCR-positive samples belonged to a single monophyletic cluster in the Indian Ocean Lineage (IOL) of the ECSA genotype of CHIKV (Figure 4). This clade is distinct from those previously identified in Western and Central Africa, suggesting that the Kassala outbreak was caused by an independent introduction of an IOL strain into the region. This might have occurred via the Middle East or Indian Subcontinent since the Kassala sequences are closely related to (i) the Henan and Shivane variants identified in China and Hong Kong, which originated in returning travellers from India/Pakistan, and (ii) a clade that includes sequences from a 2016 outbreak in Pakistan and other sequences from India and Bangladesh. Absence of the A226V mutation - a mutation associated with the capacity of the virus to infect *Ae. albopictus*^*31, 32*^ confirms *Ae. aegypti* as the most likely outbreak vector species. (Figure 5)

**Figure 5:**
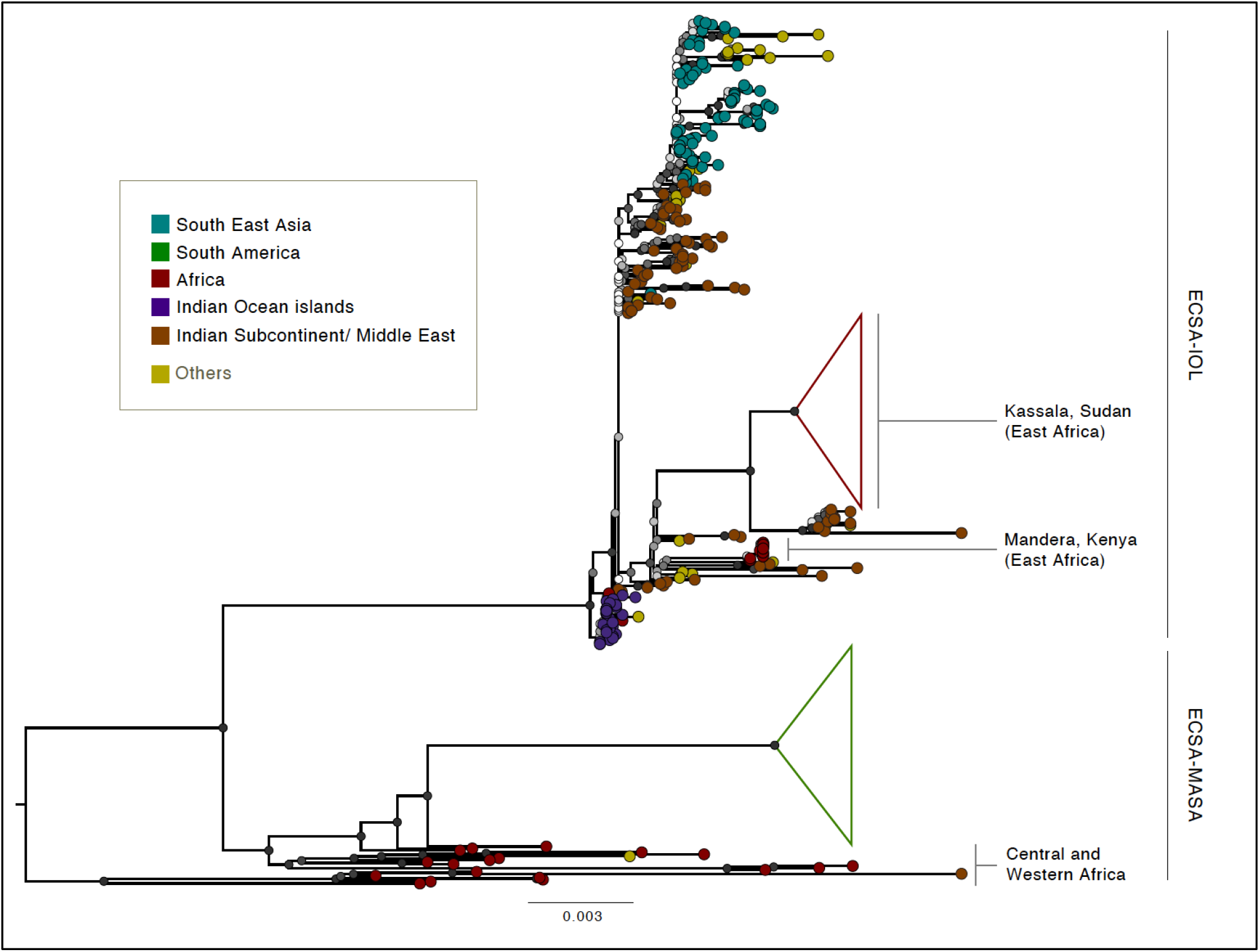
Maximum likelihood phylogenetic tree of the ECSA genotype and its two distinct lineages showing the location of the Kassala 2018/19 strain. **Legend:** Maximum likelihood phylogenetic tree of the East/Central/South African (ECSA) genotype and its two distinct lineages: the Indian Ocean Lineage (IOL) and the Middle African/ South American (MASA) lineage. The monophyletic Kassala epidemic clade groups with sequences that originate from the Middle East and India and represents a distinct lineage from other African CHIKV variants. Node support values were evaluated using 100 bootstrap replicates and are illustrated using colour at nodes (with white representing 0 and black representing 100). The 2016 Mandera outbreak also represents a possible introduction from the Indian subcontinent and appears to be an independent event from the Kassala epidemic. Both outbreaks share Ae. aegypti adaptive variant at site A226.

**Figure 5:**
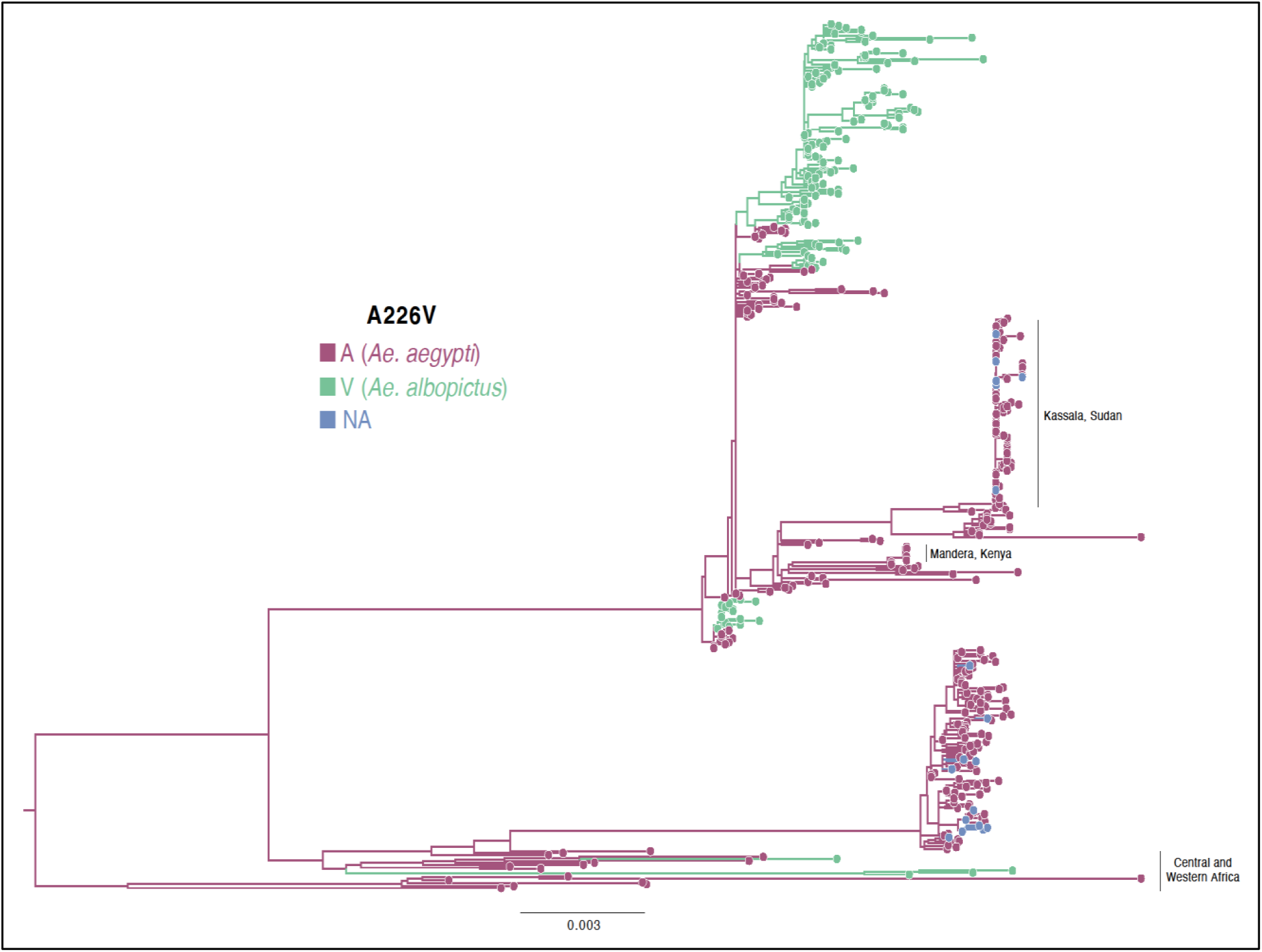
CHIKV ECSA phylogenetic tree showing the amino acid identity at site 226 of the E1 protein, associated with viral adaptation to different vector species. Viruses containing the A226 variant infect Ae. aegypti more efficiently, while viruses containing the V226 variant more efficiently infect Ae. albopictus. The evolution of this trait is mapped (through a parsimony reconstruction) on the tree in different colours.

*[Genbank accession numbers to be added]*

## DISCUSSION

Our phylogenetic analysis suggests the unprecedented Chikungunya epidemic that took place in the Eastern states of Sudan in 2018-19 was caused by an independent introduction of a CHIKV IOL virus, of the same strain responsible for the outbreaks observed worldwide during 2004-2007 and linked to more severe manifestations of CHIKV disease.

Even at the conservative estimate^33, 34^ of 487,600 cases, the epidemic that occurred in Kassala and Red Sea States is the largest outbreak of Chikungunya in Africa and the Middle East to date, and there are multiple reasons to suspect substantial under-reporting and underdiagnosis. Reasons for this include reporting through sentinel surveillance sites normally intended to raise an alert rather than capture every case, population reluctance to seek medical attention at hospitals due to cost and limited treatment options, limited diagnostic resources, lack of data collection in private clinics, and the range of CHIKV illness severity.

Made possible by the established research collaboration between the FMoH and the UK Public Health Rapid Support Team on outbreak response, this study utilised pre-approved protocols, pre-trained staff and, for the first time within Sudan, next generation sequencing technology in-country. We confirmed that CHIKV was the dominant pathogen responsible for the outbreak, that DENV was also circulating with 1 in 5 CHIKV positive patients co-infected, and importantly that there were a high frequency of falciparum malaria transmission and of CHIKV/malaria co-infection.

The cohort recruited were young (median age 27) and largely presented with non-specific febrile symptoms plus poly-articular joint pain. The substantial proportion (26%) admitted to hospital, however, reflects high frequencies of co-infection and the potential for haemodynamic disturbance in adults, and other manifestations of a severe phenotype observed in a subset of patients. As we have described, 5% of patients with CHIKV (malaria and DENV negative) had evidence of bleeding and loss of haemostasis. Two children in particular were gravely unwell with life-threatening haemorrhage associated with severe thrombocytopenia and need for blood product resuscitation, while one adult died of multi-organ failure including acute kidney injury.

Severe and fatal CHIKV infection has been reported previously during outbreaks challenging the misconception that it is only a mild self-limiting diseases.^13, 35^ Increased risk of severe disease has been suggested in extremes of age (neonates and > 65 years) and those with underlying medical problems including immunosuppression.^10, 15^ Severe disease has been associated with neurological complications such as acute flaccid paralysis and Guillain-Barré syndrome.^36-38^ Brazil, Reunion Island and India all reported high case fatality and excess deaths during their epidemics, ^12, 13, 16, 39, 40^ though fatality is still considered rare, occurring in less than 1 in 1000 cases.

These findings are consistent with the severe phenotype of CHIKV we observed in Kassala, including a one confirmed CHIKV PCR-positive death and life-threatening bleeding due to loss of haemostasis with thrombocytopenia. Additionally, the high frequency of *P*.*falciparum* infection detected underpins the importance of both malaria diagnostics and public messaging in malaria-endemic settings when chikungunya outbreaks occur.

Our study was biased towards more severe cases due to hospital-based recruitment and a low response rate among and lack of a control for participants followed up within 90-120 days. However, our findings that 63% of respondents had persistent pain three to four months after acute illness are similar to those of a case-controlled study on La Reunion Island. This study found 53% of IOL-strain CHIKV-positive participants reported twice as much pain compared to controls 12 months after their illness.^41^ Our findings were also similar to those of a meta-analysis of chronic disability across all CHIKV strains which found on average 52% of IOL-strain patients were still disturbed by symptoms at three months, compared to 39% of Asian lineage and 14% of ECSA group patients.^21^

Our finding that the only significant risk factor for contracting CHIKV was proximity to a case suggests identifying and supporting vector control measures acceptable to the community should be a priority for public health authorities seeking to prevent future CHIKV and other *Aedes*-transmitted epidemics in Sudan. Entomological research carried out by Kassala University in 2014/15 showed high *Ae. aegypti* density across the city and a high proportion of households storing water in their compounds in unprotected clay pots.^42-44^ In 2018 the situation was exacerbated by intense seasonal rainfall which displaced 50,000 residents and flooded many areas, a situation that is being seen again in 2020.

Measures such as improved water infrastructure, covered water storage containers, and increased population awareness of the importance of preventing mosquitoes from breeding in household compounds are important.^42^ Heightened vector surveillance is also needed to monitor for presence of *Ae. albopictus*, which could prompt further outbreaks of different CHIKV strains, as well as investigation of the potential role in the sylvatic cycle of the large population of Macaques monkeys which live close to residential areas.

Finally, but importantly, it is essential that State Ministries of Health throughout Sudan enhance their risk communication and public information strategies around Chikungunya to underline that severe and occasionally fatal infection exists. The epidemic we describe caused substantial health and economic burden for the affected populations. With widespread presence of *Ae. aegypti* and a similar water storage practices throughout Sudan, timely actions will be needed to prevent another large outbreak in the near future.

## Data Availability

Data files and gene sequences will be available from the LSHTM Data Compass Repository and Genbank (accession numbers - to be added shortly)

## Acknowledgements and disclaimers

The study team appreciates the collaboration of the leadership of the University of Kassala, Kassala Teaching Hospital, Kassala State Ministry of Health, Federal Ministry of Health Directorate of Health and Epidemics Control Directorate, and the National Public Health Laboratory. We also appreciate the efforts of the International Severe Acute Respiratory and Emerging Infection Consortium to create generic protocols including the Clinical Characterisation Protocol for Severe Emerging Infections from which our study was adapted.^45^ The UK Public Health Rapid Support Team is funded by UK aid from the Department of Health and Social Care and is jointly run by Public Health England and the London School of Hygiene & Tropical Medicine. The University of Oxford and King’s College London are academic partners. The views expressed in this publication are those of the authors and not necessarily those of the National Health Service, the National Institute for Health Research, or the Department of Health and Social Care.

